# Comprehensive exploration of the genetic contribution of the dopaminergic and serotonergic pathways to psychiatric disorders

**DOI:** 10.1101/2020.06.30.20143404

**Authors:** Judit Cabana-Domínguez, Bàrbara Torrico, Andreas Reif, Noèlia Fernàndez-Castillo, Bru Cormand

## Abstract

Psychiatric disorders are highly prevalent and display considerable clinical and genetic overlap. Dopaminergic and serotonergic neurotransmission have been shown to have an important role in many psychiatric disorders. Here we aim to assess the genetic contribution of these systems to eight psychiatric disorders (ADHD, ANO, ASD, BIP, MD, OCD, SCZ and TS) using publicly available GWAS analyses performed by the Psychiatric Genomics Consortium. To do so, we elaborated four different gene sets using the Gene Ontology and KEGG pathways tools: two ‘wide’ selections for dopamine (DA) and for serotonin (SERT), and two ‘core’ selections for the same systems. At the gene level, we found 67 genes from the DA and/or SERT gene sets significantly associated with one of the studied disorders, and 12 of them were associated with two different disorders. Gene-set analysis revealed significant associations for ADHD and ASD with the wide DA gene set, for BIP with the wide SERT gene set, and for MD with both the core DA set and the core SERT set. Interestingly, interrogation of the cross-disorder GWAS meta-analysis displayed association with the wide DA gene set. To our knowledge, this is the first time that these two neurotransmitter systems have systematically been inspected in these disorders. Our results support a cross-disorder contribution of dopaminergic and serotonergic systems in several psychiatric conditions.

## INTRODUCTION

Psychiatric disorders represent a major health problem, affecting 29.2% of the worldwide population at some point during their lifetime [1], and are associated with a considerable distress and functional impairment [2]. They constitute a set of complex traits that result from the interaction of genetic and environmental risk factors, with a heritability ranging from 40 to 80%, depending on the disorder, as estimated by twin studies [3].

Interestingly, psychiatric disorders display considerable clinical overlap among them [4–6], and the presence of comorbidity is associated with an increased severity and difficulty of treatment. Recent studies have shown strong genetic correlations among psychiatric phenotypes [7,8], and the latest genome-wide association study (GWAS) meta-analysis conducted on eight psychiatric disorders (attention-deficit hyperactivity disorder (ADHD), anorexia nervosa (ANO), autism spectrum disorder (ASD), bipolar disorder (BIP), major depression (MD), obsessive-compulsive disorder (OCD), schizophrenia (SCZ) and Tourette’s syndrome (TS)) found that 75% of the LD-independent associated regions (109 out of 146) were associated with more than one disorder [9]. These results suggest that the high levels of comorbidity found in psychiatric disorders may be explained, at least in part, by shared genetic risk factors, supporting the existence of a set of genes that confer relatively broad liability to psychiatric disorders [9].

Dopamine (DA) and serotonin (SERT) are two important neurotransmitters that participate in the regulation of a wide range of essential functions of the organism (e.g. motor control, cognition, motivation, regulation of emotions or reward), and have been related to the physiopathology and treatment of many psychiatric disorders.

Dopaminergic dysfunction has been described in ADHD, ASD, OCD, TS, SCZ, mood disorders and substance use disorders (SUD) among others [10]. For example, the positive symptoms of SCZ seem to be associated with hyperdopaminergic neurotransmission, especially in the mesolimbic system, while the negative symptoms and cognitive deficits might be caused by hypodopaminergic activity in the mesocortical pathway [11]. Therefore, most of the antipsychotic treatments block the dopamine D2 receptor (e.g. chlorpromazine and haloperidol), and some of them are combined with serotonin 5-HT2A receptor antagonists (e.g. clozapine and risperidone). Moreover, several studies have reported a reduction of dopaminergic receptors density in several brain regions of ADHD patients [12], which is in agreement with the mechanism of action of the ADHD treatments, like methylphenidate or amphetamine, that enhance dopamine transmission in prefrontal cortex [13].

Regarding the serotonergic system, there is strong evidence supporting its role in MD and in other mood disorders based on available pharmacological treatments. The serotonin hypothesis of depression postulates that reduced serotonin signaling is a risk factor in its etiology [14,15], what is supported by the most effective antidepressant treatment, based on the use of serotonin selective reuptake inhibitors (SSRIs), which inhibit the serotonin transporter increasing the extracellular levels of the neurotransmitter [16]. However, the causal mechanisms between low serotonin signaling and depression are still unknown. The role of the serotonergic system in other psychiatric disorders like anxiety, SCZ, ADHD or ASD is still unclear and further studies are needed [17]. Even so, all these psychiatric conditions seem to be related to serotonin dysfunction, and many psychotropic drugs interfere more or less directly with this system [18].

Dopaminergic and serotonergic neurotransmission have been widely investigated through candidate-gene association studies in many psychiatric disorders, which have mainly assessed genetic variants in core genes encoding DA and SERT transporters, receptors and metabolizers [19–22]. One of the most studied genes is *SLC6A3*, encoding the dopaminergic transporter (DAT), with multiple variants mapping at this locus, including rare variants, copy number variations (CNVs), variable number of tandem repeats (VNTRs) and single nucleotide polymorphisms (SNPs) that have been found associated with several psychiatric conditions [23]. Although genetic variants in the serotonin transporter (SERT), encoded by *SLC6A4*, have also been extensively studied in psychiatric behavioral genetics, these different studies failed to obtain consistent results [24]. Interestingly, the outcome of genome-wide association studies (GWAS) evidenced that the classical serotonergic and dopaminergic candidate genes, like transporters (*SLC6A3* and *SLC6A4*) and receptors are not significantly associated with any psychiatric disorder (https://www.ebi.ac.uk/gwas/home), with one exception, *DRD2*, that was found associated with both MD and SCZ [25–27].

In this study, we systematically explored the contribution of common variants in genes involved in dopaminergic and serotonergic neurotransmission in eight psychiatric disorders studied individually and in combination, using GWAS data from the Psychiatric Genomics Consortium (PGC).

## MATERIALS AND METHODS

### DA and SERT gene selection

We first obtained two core gene sets through manual curation (DA core with 13 genes, and SERT core with 20 genes), containing the well-known dopaminergic or serotonergic genes (neurotransmitter receptors, transporters and enzymes involved in their synthesis or degradation) (Figure 1A).

**Figure 1.**
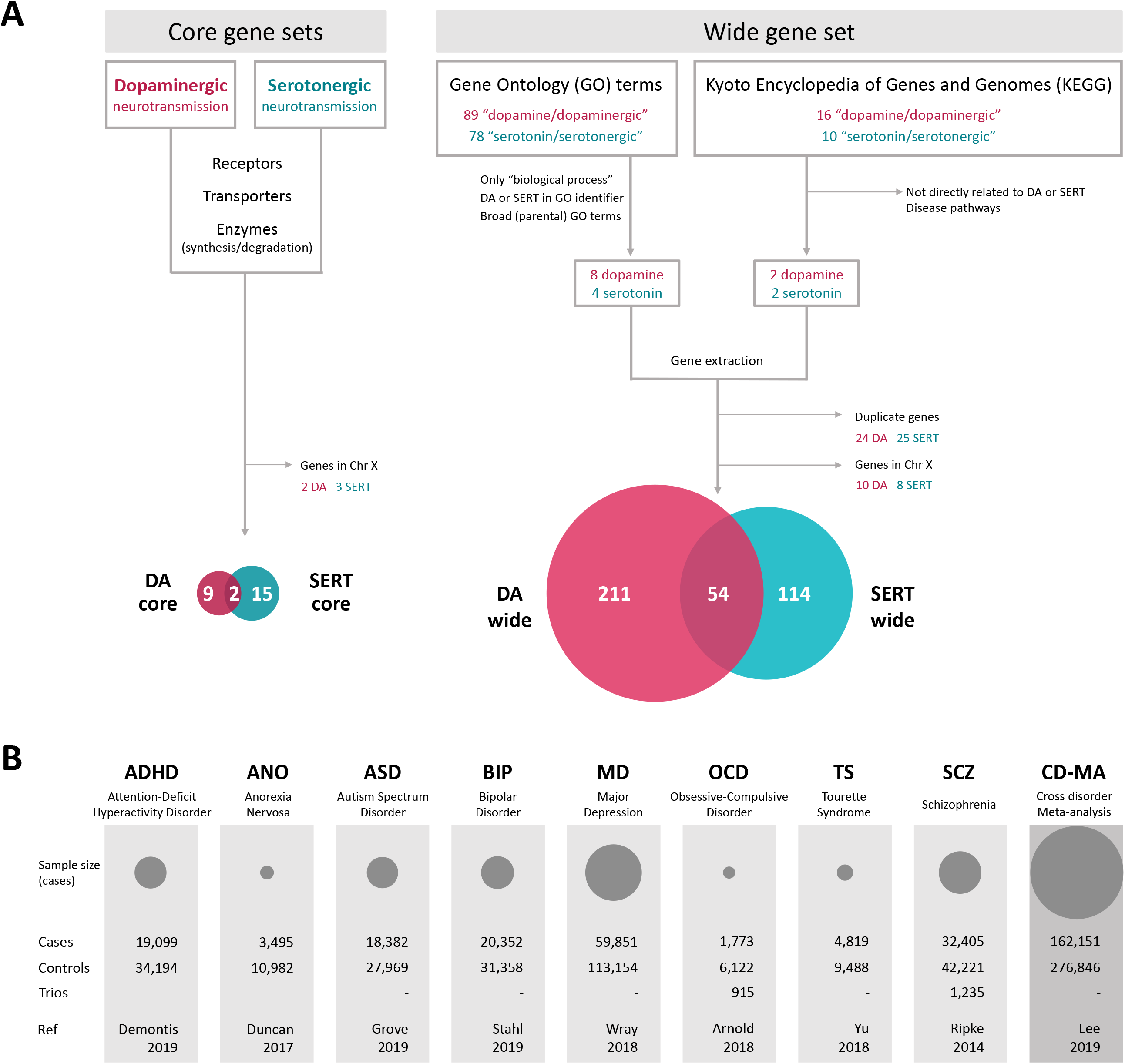
(A) Strategy for the selection of the serotonin (SERT) and dopamine (DA) gene sets using Gene Ontology and KEGG’s pathways for the wide sets and manual curation for the core sets. All genes of the core gene sets are also included in their corresponding wide gene set. (B) Description of samples used in this study. The area of the circles is proportional to the sample size of the disorder.

For a wide selection of genes encoding proteins involved in dopamine and serotonin neurotransmission and signaling pathways we used two main databases, GO and KEGG. We queried the GO database (Gene Ontology Consortium, http://www.geneontology.org/, September 2016) by the search terms “dopamine” or “serotonin/serotonergic”. The resulting list of GO terms (89 for dopamine and 78 for serotonin/serotonergic) was filtered to keep only those of “biological process” ontology source and with presence of the search term as part of the GO identifier. Then, we examined the hierarchical tree structure of the GO database to filter out the most specific terms (child terms) that were contained within broader ones (parental terms). The final GO terms selection included: “dopamine transport” (GO:0015872), “dopamine receptor signaling pathway” (GO:0007212), “dopamine receptor binding” (GO:0050780), “synaptic transmission, dopaminergic” (GO:0001963), “dopaminergic neuron axon guidance” (GO:0036514), “dopaminergic neuron differentiation” (GO:0071542), “response to dopamine” (GO:1903350), “dopamine metabolic process” (GO:0042417), “serotonin receptor signaling pathway” (GO:0007210), “serotonin transport” (GO:0006837), “serotonin metabolic process” (GO:0042428) and “serotonin production involved in inflammatory response” (GO:0002351). For the exploration of KEGG pathways (https://www.genome.jp/kegg/pathway.html, September 2016) we followed a similar procedure, using the terms “dopamine” or “serotonin” as keywords, and only those pathways more tightly related to neurotransmitters’ function were kept: “dopaminergic synapse” (hsa04728) and “tyrosine metabolism” (hsa00350), and “serotonergic synapse” (hsa04726) and “tryptophan metabolism” (hsa00380). The human entries obtained were then filtered for more inclusive and non-disease pathways.

We elaborated the dopaminergic (DA wide, 275 genes) and serotonergic (SERT wide, 176 genes) gene sets with human genes belonging to the final GO categories plus the selected KEGG pathways. We confirmed that these two lists contained the corresponding core genes. As the summary statistics used in our analyses did not include the X chromosome we filtered out those genes from our gene sets. In the case of DA genes, 10 genes were removed: 2 core genes (*MAOA* and *MAOB*) and 8 other genes from the wide set (*ATP7A, AGTR2, FLNA, GPR50, GRIA3, HPRT1, PPP2R3B* and *VEGFD)*. And 8 genes were excluded for SERT: 3 core genes (*MAOA, MAOB* and *HTR2C*) and 5 other genes from the wide set (*ATP7A, ARAF, ASMT, CACNA1F* and *GPM6B*) (Figure 1A).

Finally, four gene lists were obtained: i) DA core (11 genes); ii) DA wide (265 genes); iii) SERT core (17 genes); and iv) SERT wide (168 genes) (Figure 1A).

Gene symbols were converted to Entrez gene ID with the DAVID Gene ID Conversion Tool (https://david.ncifcrf.gov/conversion.jsp) for downstream analysis.

### Data used

We inspected the four DA and SERT gene sets in eight psychiatric disorders and in the meta-analysis of all of them (cross-disorder meta-analysis; CD-MA) [9] using publicly available GWAS summary statistics of attention-deficit hyperactivity disorder (ADHD) [28], anorexia nervosa (ANO) [29], autism spectrum disorder (ASD) [30], bipolar disorder (BIP) [31], major depression (MD) [32], obsessive-compulsive disorder (OCD) [33], schizophrenia (SCZ) [26] and Tourette’s syndrome (TS) [34]. All of them were downloaded from the Psychiatric Genomics Consortium (PGC) webpage (https://www.med.unc.edu/pgc/results-and-downloads/) (Figure 1B). The MD and CD-MA summary statistics do not include 23andMe data used by the PGC in the MD GWAS.

### Gene-based and gene-set association analyses

Gene-based and gene-set association analyses of each phenotype were performed with MAGMA 1.06 [35] using the summary statistics from each individual GWAS meta-analysis and also from the cross-disorder GWAS meta-analysis. For the gene-based analysis the SNP-wise mean model was used, in which the test statistic used was the sum of −log(SNP p-value) for SNPs located within the transcribed region (defined on NCBI 37.3 gene definitions), with a 0 Kb window around genes. MAGMA accounts for gene size, number of SNPs in a gene and linkage disequilibrium between markers, using as a reference panel the European ancestry samples from the 1000 Genomes Project, phase 3 [36]. The resulting p-values were corrected for multiple testing using False Discovery Rate (5% FDR).

Based on the gene-based p-values we analysed the four sets of genes described above (DA wide, DA core, SERT wide and SERT core). MAGMA applies a competitive test to analyse whether the genes of a gene set are more strongly associated with the trait than other genes, while correcting for a series of confounding effects such as gene length and size of the gene set. In our analyses, only genes on autosomes were included. Multiple testing corrections were performed for each disorder separately. As the Bonferroni correction is quite conservative for gene sets strongly overlapped, we used an empirical multiple testing correction implemented in MAGMA, based on a permutation procedure.

### Gene-based Manhattan plot

Results of the gene-based analysis of the cross-disorder meta-analysis were plotted in R using the package “qqman” [37].

### MetaXcan analyses

We considered all the SNPs located in each DA and SERT gene to infer whether the genetically determined expression of each of those genes correlates with the phenotypes considered in the study. These analyses were carried out on MetaXcan [38] using the summary statistics of each disorder. Prediction models were constructed considering SNPs located within ± 1Mb from the transcription start site (TSS) of each gene and were trained with RNA-Seq data of 13 GTEx (release V7) brain tissues [39]. The SNP covariance matrices were generated using the 1000 Genomes Project Phase 3 [36] EUR genotypes of the prediction model SNPs. For each brain tissue, the threshold for significance was calculated using the Bonferroni correction for multiple testing.

## RESULTS

Dopaminergic and serotonergic neurotransmission have been pointed out as involved in many psychiatric disorders. In the present study we aimed to assess the contribution of these two monoaminergic systems to eight different psychiatric disorders (ADHD, ANO, ASD, BIP, MD, OCD, SCZ and TS) as well as to the meta-analysis of all them (CD-MA). For that purpose, we elaborated four different gene sets (DA wide, 265 genes; SERT wide, 168 genes; DA core, 11 genes; and SERT core, 17 genes; Figure 1A and Supplementary Table S1) that were subsequently interrogated in the summary statistics from nine case-control GWAS meta-analyses (Figure 1B). One gene from DA wide (*KIF5C*) and two from SERT wide (*OR11H7* and *OR10J6P*) gene sets were not present in any of the datasets inspected. At the gene-wide level, we found 67 genes from DA and/or SERT gene sets significantly associated (5% FDR) with at least one of the studied disorders (Supplementary Table S2). Interestingly, twelve of these genes were associated with two different disorders, eight of them with SCZ and BIP (Figure 2 and Supplementary Table S2). Eleven out of these twelve genes showed also a significant association with the CD-MA phenotype, *DRD2* from the DA core gene set among them. Interestingly, five out of these twelve genes (*CACNA1C, CACNA1D, GNAS, GRIN2A* and *ITR3*) belong to both the DA and SERT gene sets, highlighting the importance of those genes that are involved in two monoamine pathways (Supplementary Table S2).

**Figure 2.**
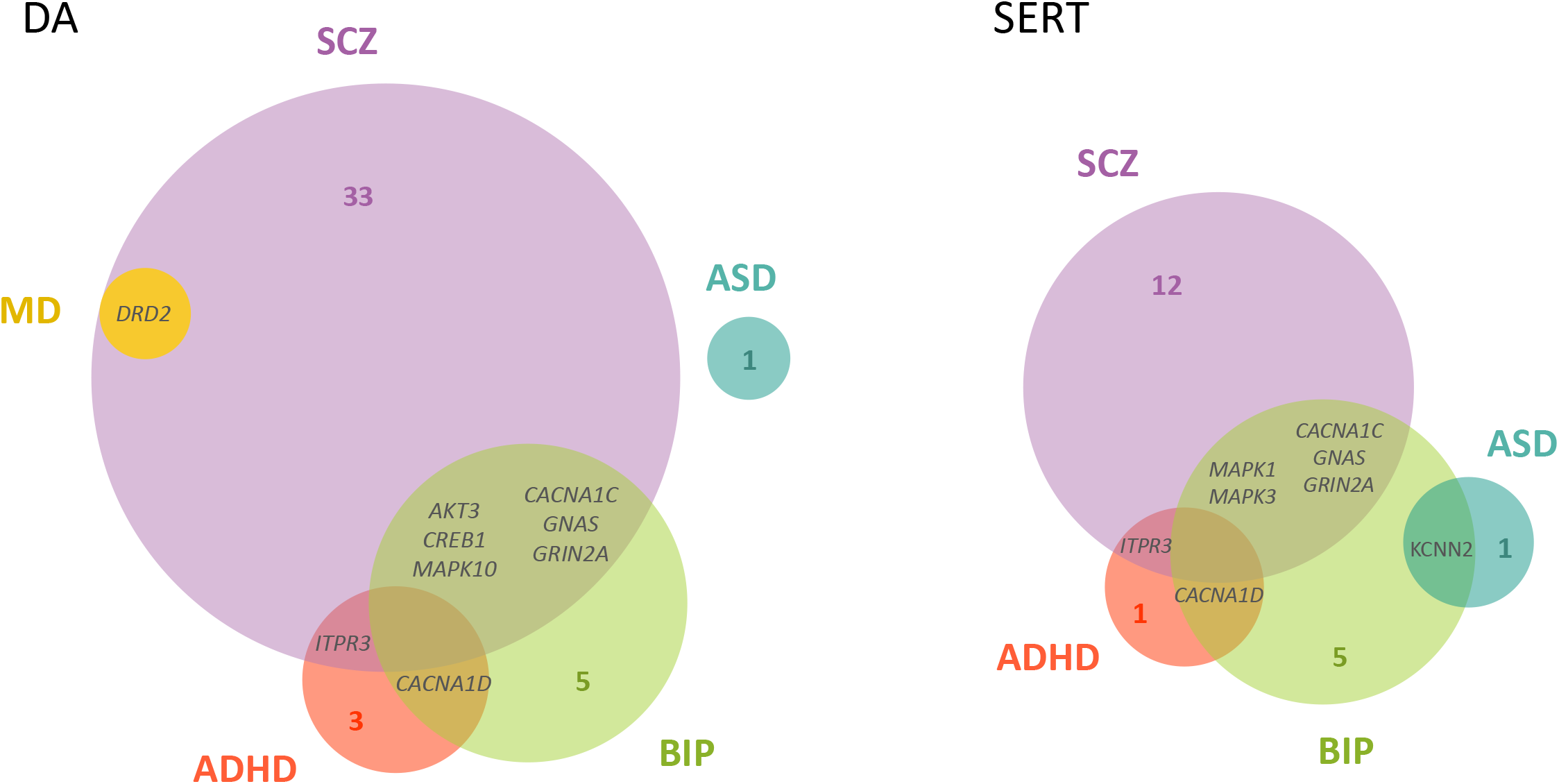
Venn diagram of the significantly associated DA and SERT genes across the studied disorders (5% False Discovery Rate). Gene names are only provided for those genes significantly associated with more than one disorder (overlapping genes). All the genes overlapping across phenotypes are significantly associated with the phenotype in the cross-disorder meta-analysis, except for GNAS. The area of circles is proportional to the number of significantly associated genes in each disorder. DA: dopaminergic gene set; SERT: serotonergic gene set. ADHD: Attention-Deficit/Hyperactivity Disorder; ASD: Autism Spectrum Disorder; BIP: Bipolar Disorder; MD: Major Depression; SCZ: Schizophrenia.

Then, we plotted the gene-based results of the cross-disorder meta-analysis in a Manhattan plot and highlighted the DA core and SERT core genes, to visualize the performance of these genes in the combination of all the psychiatric conditions considered in the study (Figure 3A). As shown in the figure, only the *DRD2* gene overcomes the Bonferroni correction for multiple testing, and *HTR6* surpasses 5% FDR. Then, we repeated the analysis by highlighting the DA and SERT wide genes and found 10 genes with associations that overcome the Bonferroni correction and 32 additional genes overcoming 5% FDR (Figure 3B). Among them, *CACNA1C*, present in both DA and SERT gene lists, was the one showing the strongest association in the cross-disorder meta-analysis (raw *P*=8.9E-14, *P*adj=7.8E-11).

**Figure 3.**
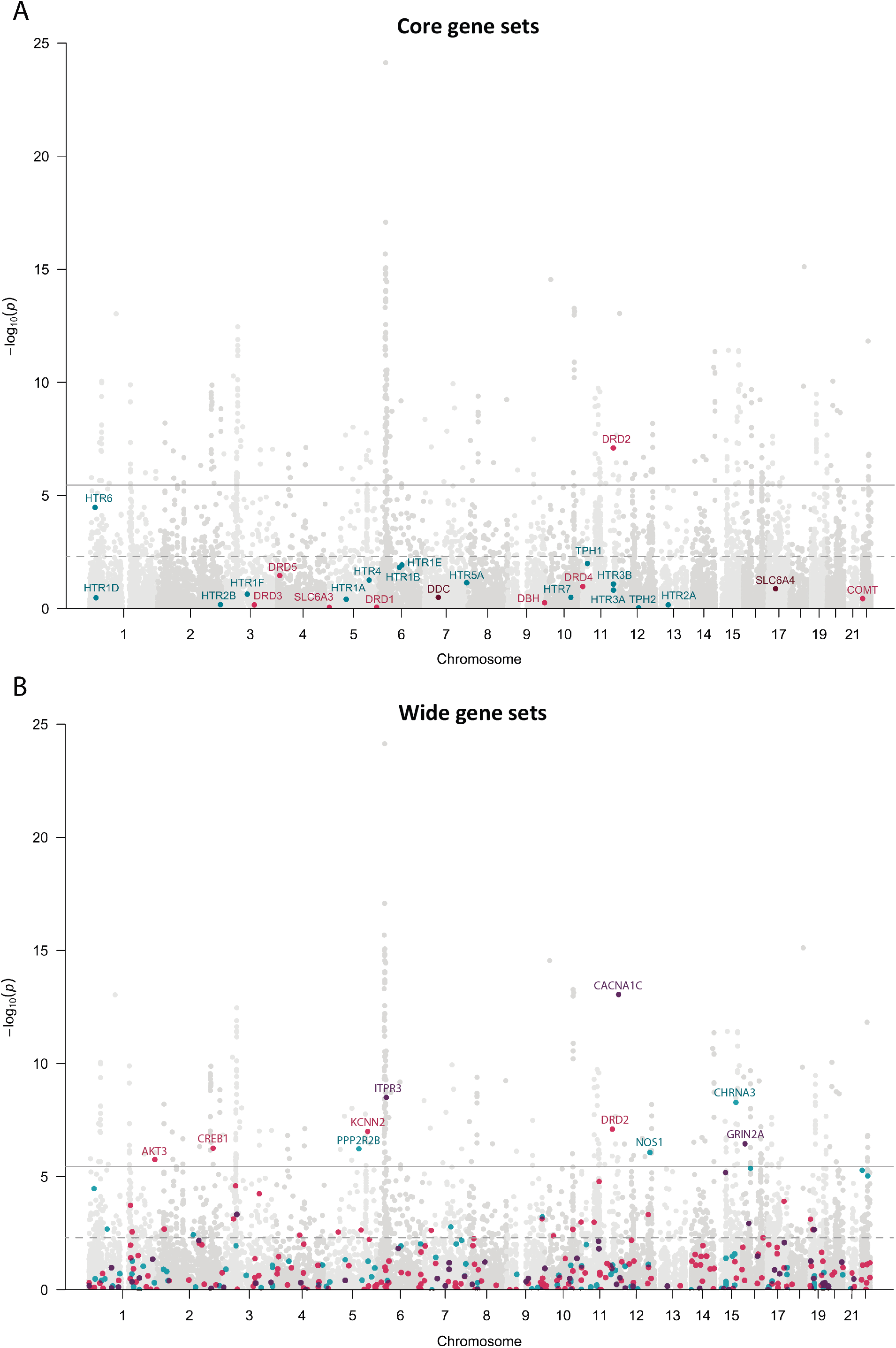
Manhattan plots of the gene-based p-values in the cross-disorder meta-analysis. Represented A) DA core and SERT core genes, and B) DA wide and SERT wide gene sets. Colored dots correspond to DA genes in pink, SERT genes in blue and overlapping genes in purple. Continuous line: threshold for Bonferroni significance (P = 2.8E-06). Discontinuous line: threshold for 5% False Discovery Rate (P = 4.9E-03).

In addition, to identify genes which expression correlates with the phenotypes included in the study, we performed MetaXcan analysis, considering only those phenotypes that show significant association with DA or SERT genes (ADHD, ASD, MD, SCZ and CD-MA). Taking in account the genes associated at gene level with a given disorder, we found that the expression of *CTNNB1* significantly correlates with ADHD in the amygdala (*P*=8.9E-04, Z=-3.3246), *ATF6B* with SCZ in the spinal cord (*P*=1.6E-06, Z=4.7925) and *DNM1* and *ITPR3* with the phenotype in the CD-MA in the cortex and caudate, respectively (*P*=2.6E-04, Z=3.6516 and *P*=1.8E-05, Z=-4.2860) (Table 1 and Supplementary Tables S3-S15). Interestingly, the expression of the *DNM1* gene in the cortex also correlates with ADHD (*P*=8.5E-03, Z=2.6308), ASD (*P*=5.9E-03, Z=2.7524), MD (*P*=3.0E-03, Z=2.9630) and SCZ (*P*=0.0312, Z=2.1549) in the same direction, and this gene was found nominally associated with all these disorders in the gene-based analysis (raw *P*_ADHD_ =2.5E-04, raw *P*_ASD_ =3.6E-03, raw *P*_MD_ =1.5E-03 and raw *P*_SCZ_ =0.0156). It is important to note that for most DA and SERT genes (ranging from 77% to 93.6% depending on the tissue and disorder) we could not retrieve information from MetaXcan because it only provides data about those gene models where the predictive power is good enough.

**Table 1.** MetaXcan prediction of gene expression effects on the studied disorders for multiple brain tissues.

| Gene symbol | entrezID | Gene sets |  | Brain tissue<sup>&</sup> | Disorder | Z-score | p-value | N SNPs in model | N SNPs used | Predicted R<sup>2</sup> |
| --- | --- | --- | --- | --- | --- | --- | --- | --- | --- | --- |
| *ATF6B* | 1388 | DA |  | Spinal cord | SCZ | 4.7924 | **1.6E-06** | 11 | 5 | 0.1466 |
| *CACNA1C* | 775 | DA | SERT | Cerebellum | CD-MA | -3.0166 | 2.6E-03 | 51 | 41 | 0.2435 |
|  |  |  |  |  | SCZ | -2.4867 | 0.0129 | 51 | 50 | 0.2435 |
| *CACNA1D* | 776 | DA | SERT | Cerebellar hemisphere | ADHD | -2.2252 | 0.0261 | 107 | 89 | 0.0988 |
| *CTNNB1* | 1499 | DA |  | Amygdala | ADHD | -3.3246 | **8.9E-04** | 36 | 35 | 0.1692 |
|  |  |  |  | Caudate | ADHD | -2.8011 | 5.1E-03 | 12 | 12 | 0.0789 |
|  |  |  |  | Putamen | ADHD | -3.0637 | 2.2E-03 | 28 | 27 | 0.1740 |
|  |  |  |  | Spinal cord | ADHD | -2.2299 | 0.0258 | 47 | 32 | 0.1377 |
| *CYP2D6* | 1565 |  | SERT | Amygdala | CD-MA | 2.5247 | 0.1158 | 15 | 15 | 0.2419 |
|  |  |  |  |  | SCZ | 2.3181 | 0.0204 | 15 | 15 | 0.2419 |
| *DNM1* | 1759 | DA |  | Cortex | ADHD | 2.6308 | 8.5E-03 | 3 | 3 | 0.0502 |
|  |  |  |  |  | ASD | 2.7524 | 5.9E-03 | 3 | 3 | 0.0502 |
|  |  |  |  |  | CD-MA | 3.6516 | **2.6E-04** | 3 | 3 | 0.0502 |
|  |  |  |  |  | MD | 2.9630 | 3.0E-03 | 3 | 3 | 0.0502 |
|  |  |  |  |  | SCZ | 2.1548 | 0.0312 | 3 | 3 | 0.0502 |
| *FLOT1* | 10211 | DA |  | Cerebellar hemisphere | SCZ | -2.2126 | 0.0269 | 79 | 5 | 0.1378 |
| *GCH1* | 2643 | DA |  | Cerebellum | SCZ | -1.1404 | 0.0490 | 103 | 102 | 0.0552 |
| *GNB2* | 2783 |  | SERT | Spinal cord | CD-MA | -2.8822 | 3.9E-03 | 31 | 29 | 0.0579 |
| *GRIN2A* | 2903 | DA | SERT | Spinal cord | BIP | -2.6559 | 7.9E-03 | 3 | 3 | 0.2636 |
| *HTR6* | 3362 |  | SERT* | Anterior cingulate | BIP | 2.4006 | 0.0164 | 58 | 58 | 0.0968 |
|  |  |  |  |  | CD-MA | 2.4828 | 0.1303 | 58 | 49 | 0.0968 |
| *ITPR3* | 3710 | DA | SERT | Caudate | CD-MA | -4.2860 | **1.8E-05** | 4 | 4 | 0.0867 |
| *MAPK3* | 5595 |  | SERT | Hippocampus | SCZ | 2.0050 | 0.0449 | 17 | 17 | 0.0734 |
| *MAPK10* | 5602 | DA |  | Anterior cingulate | BIP | 2.3373 | 0.0194 | 15 | 15 | 0.1481 |
|  |  |  |  |  | CD-MA | 2.8561 | 0.0344 | 15 | 15 | 0.1481 |
|  |  |  |  | Caudate | BIP | 2.0394 | 0.0414 | 39 | 39 | 0.1546 |
|  |  |  |  |  | CD-MA | 2.1152 | 0.0344 | 39 | 32 | 0.1546 |
|  |  |  |  | Cortex | BIP | 2.6829 | 7.3E-03 | 28 | 28 | 0.0880 |
|  |  |  |  |  | CD-MA | 2.6987 | 7.0E-03 | 28 | 22 | 0.0880 |
|  |  |  |  | Nucleus accumbens | BIP | 2.1524 | 0.0314 | 35 | 35 | 0.1187 |
|  |  |  |  |  | CD-MA | 2.3767 | 0.0175 | 35 | 33 | 0.1187 |

<sup>&</sup>Prediction models were only available for some tissues and genes; Z-score: Number of standard deviations change in gene expression; p-value: Significance of the association between predicted expression levels and the disorder; N SNPs in model: Number of SNPs used in the training of prediction models for each gene; N SNPs used: Number of SNPs used from the corresponding GWAS summary statistics; Predicted R<sup>2</sup>: Correlation between the predicted and observed gene expression during prediction model training; DA: Wide dopaminergic gene set; SERT: Wide serotonergic gene set. \* Gene present in SERT core gene set. ADHD: Attention-Deficit/Hyperactivity Disorder; ASD: Autism Spectrum Disorder; BIP: Bipolar Disorder; MD: Major Depression; SCZ: Schizophrenia; CD-MA: Cross-Disorder meta-analysis. In bold: Significant p-values overcoming Bonferroni correction for multiple testing.

Finally, we performed a gene-set analysis that revealed interesting significant associations. The DA wide gene set was significantly associated with ADHD (*P*_perm_=0.023) and ASD (*P*_perm_=0.025), and the SERT wide gene set with BIP (*P*_perm_=0.015). Besides, both the DA core (*P*_perm_=7.1E-03) and the SERT core (*P*_perm_=0.021) gene sets were significantly associated with MD (Figure 4 and Supplementary Table S16). Interrogation of the cross-disorder GWAS meta-analysis displayed association with the DA wide gene set (*P*_perm_=0.044). It should be mentioned that the DA wide gene set was nominally associated also with BIP, TS and SCZ, in line with the results obtained in the cross-disorder analysis for this neurotransmitter system (Figure 4). No significant results were found for ANO, OCD and TS neither in the gene-based nor in the gene-set analyses after multiple testing correction, maybe due to limited sample sizes (number of patients below 5000; Figure 1B).

**Figure 4.**
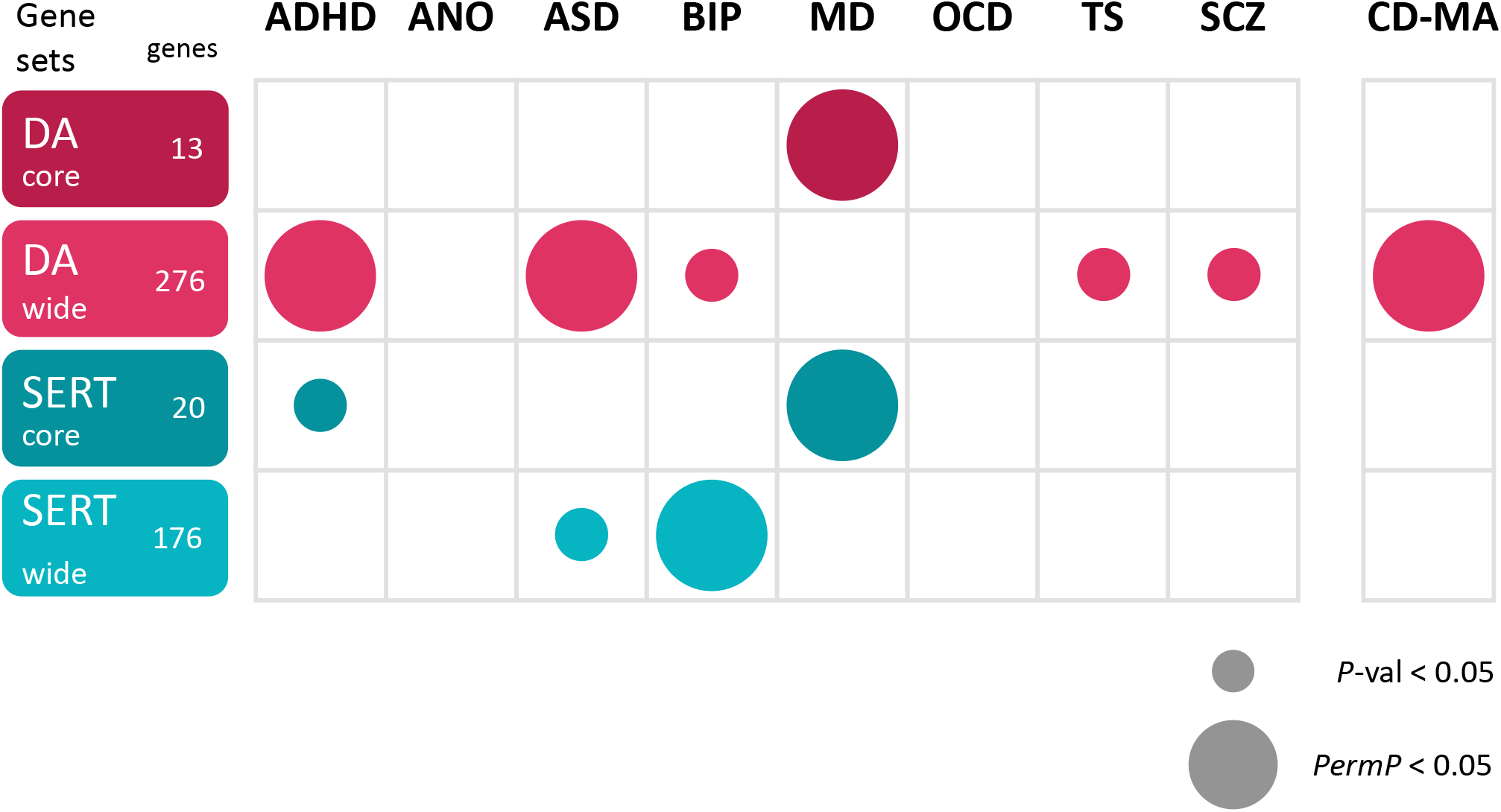
Results of the MAGMA gene-set association analysis on every phenotype under study. A small circle represents nominally significant p-value, a big circle represents significant p-value after the empirical multiple testing correction implemented in MAGMA. DA: dopaminergic gene set; SERT: serotonergic gene set. ADHD: Attention-Deficit/Hyperactivity Disorder; ANO: Anorexia Nervosa; ASD: Autism Spectrum Disorder; BIP: Bipolar Disorder; MD: Major Depression; OCD: Obsessive-Compulsive Disorder; TS: Tourette’s Syndrome; SCZ: Schizophrenia; CD-MA: Cross-Disorder.

## DISCUSSION

Dopaminergic (DA) and serotonergic (SERT) functions represent good candidates for psychiatric disorders, mainly on the basis of the effectiveness of several pharmacological treatments and their target molecules [10,23,40–42]. Common genetic variation of genes encoding key participants of these two neurotransmission systems (receptors, transporters and enzymes) has been broadly studied in psychiatric disorders, mainly through candidate-gene association studies, with several positive findings [23,43–45]. However, except for a few cases [25–27], these hits have not been replicated by GWAS, which raises questions on how alterations in these two neurotransmitter systems can actually be causally related to psychiatric conditions, but also on the value of genome-wide significance for functional variation [46].

To our knowledge, this is the first systematic study of the DA and SERT neurotransmitter systems in eight psychiatric disorders (ADHD, ANO, ASD, BIP, MD, OCD, SCZ and TS) using data from GWAS meta-analyses that include large number of samples. Here, we assess the contribution of genetic variation not only of genes encoding the core participants of DA and SERT functions, but also of a comprehensive list of genes involved in DA and SERT neurotransmission obtained by a systematic database search.

Our results show a contribution to psychiatric disorders of both dopaminergic and serotonergic systems at genetic level. We found several DA and SERT genes associated with some psychiatric disorders, twelve of them with two conditions, and also in the cross-disorder meta-analysis, which underscores the relevance of genetic risk factors in these genes for psychiatric disorders. Among the associated genes we identified the DA core gene coding for the dopamine receptor D2, *DRD2*, which has been widely studied as a candidate for different psychiatric disorders with some positive findings [47–51], especially in SCZ (recently meta-analyzed by [52]). The association between *DRD2* and SCZ observed through candidate-gene studies has subsequently been confirmed through GWAS [26]. Interestingly, a recent GWAS identified association of polymorphisms in *DRD2* with depressive symptoms, neuroticism and different traits of the well-being spectrum [53,54]. These findings are in agreement with ours, obtaining associations of *DRD2* with SCZ and MD (Figure 2 and Supplementary Table S2). Another interesting gene that encodes the calcium voltage-gated channel subunit alpha1 C (*CACNA1C*), present in both the DA and SERT wide gene lists and found associated by us with BIP and with SCZ (disorders with a very high genetic correlation, rg=0.7 [9]), displayed the strongest association in the cross-disorder meta-analysis. Noteworthy, *CACNA1C* polymorphisms have been consistently associated with BIP and SCZ, among other psychiatric disorders [55,56]. Mutations in this gene have been related with Timothy Syndrome, a monogenic disorder that affects multiple organs and presents with cognitive impairment, major developmental delays, and autism-like behaviours [57]. We also identified the potassium calcium-activated channel subfamily N member 2 (*KCNN2*) gene, from the SERT wide gene set, associated with BIP and with ASD, as previously described in two studies [30,31].

Interestingly, two genes emerged from the analysis of gene expression using MetaXcan. First, the gene for inositol 1,4,5-trisphosphate receptor type 3 (*ITPR3*), found associated with ADHD, SCZ and the CD-MA at gene level in our study, which expression would be altered in the caudate of patients from the CD-MA. The caudate nucleus is a crucial component of the ventral striatum, part of the basal ganglia that control motor, cognitive control, motivational and emotional processing [58], functions involved in the physiopathology of many psychiatric conditions. Several studies have found alterations both in the volume of the caudate nucleus and in its functional connections with other brain regions in many psychiatric disorders [59–61]. Interestingly, the *ITPR3* gene has been associated to type 1 diabetes [62], which curiously shows a correlation with risk of ADHD in the offspring [63–65]. The other gene that was pointed by this analysis was dynamin 1 (*DNM1*), which expression significantly correlates not only with the CD-MA, but also with ADHD, ASD, MD and SCZ, in all cases in the cortex and in the same direction. Rare variants in *DNM1* have been identified in patients with a mendelian phenotype, epileptic encephalopathy [66], and in some cases of intellectual disability with seizures [67].

In the cross-disorder meta-analysis study [9], the eight psychiatric disorders used were classified in three groups using exploratory factor analysis (EFA): disorders characterized by compulsive/perfectionistic behaviors (AN, OCD and TS), mood and psychotic disorders (MD, BIP and SCZ), and early-onset neurodevelopmental disorders (ASD, ADHD, TS, as well as MD). Our results are in line with this classification, as we identified association of the serotonergic gene sets with BIP and MD, included in the same group, whereas the dopaminergic set was associated with ADHD, ASD and MD, the three disorders conforming the third factor. Interestingly, MD showed association with both the SERT and the DA core gene sets and, in parallel, it represents a shared disorder between two different factors of the CD-MA.

Although SERT dysfunction has been proposed to be the common denominator in a wide range of neuropsychiatric illnesses [66], our gene-set results link this neurotransmitter system only with BIP (wide gene set) and with MD (core gene set). SERT signaling has been associated with concrete alterations in the intrinsic activity of the brain: an increase of the default-mode network and a decrease of the sensorimotor network. Interestingly, MD is characterized by an increase of internally focused thoughts and an inhibited psychomotor behavior and affectivity, an imbalance that has been also detected in BIP [66], but not in SCZ, the third of this genetically-correlated group of psychiatric disorders [9].

Gene-set analysis of the DA system showed association of the wide selection with ADHD and with ASD, two disorders that co-occur frequently [68] and with a high genetic correlation (rg= 0.37) [9]. Association of dopaminergic genes with these psychiatric conditions, especially the DA core genes, has been broadly studied using different strategies, including candidate-gene association studies or exome/genome sequencing [69–72]. Besides, the DA core gene set was significantly associated with MD. Noteworthy, many of the depressive symptoms have been consistently associated with dopaminergic dysfunction [73,74]. Interestingly, the cross-disorder meta-analysis displayed association with the DA wide gene set, underscoring the importance that this neurotransmitter has in many psychiatric disorders [75,76].

Some strengths and limitations of our study should be discussed. We performed a systematic study of the dopaminergic and serotonergic neurotransmitter systems in eight psychiatric disorders and the meta-analysis of all of them. At gene level, we identified twelve genes significantly associated with several disorders. However, none of them was associated in more than two disorders, being BIP and/or SCZ one of these disorders in most cases (10 out to 12). This brings up the issue that these two disorders could drive some of the results obtained because of their large sample sizes. In addition, DA and SERT neurotransmission are functionally interconnected and some genes participate in both pathways (about 20% of genes in the DA gene sets are also in SERT, and 32% the other way round). This could have some impact in the results obtained in the gene-set analysis. In addition, is important to note that these pathways are still not very well defined and we might be missing information from some genes. On the other hand, as MetaXcan only provides information about those gene models where the predictive power is good enough, we could not retrieve information from most DA and SERT genes. Finally, it is important to note that no significant results were found for ANO, OCD and TS neither in the gene-based nor in the gene-set analyses, maybe due to limited sample sizes. In these disorders, further studies should be done with larger samples.

To our knowledge, this is the first systematic genetic study of the dopaminergic and serotonergic neurotransmitter systems in eight psychiatric disorders (ADHD, ANO, ASD, BIP, MD, OCD, SCZ and TS) and in the meta-analysis of all of them. At gene level, we identified association of 67 DA and/or SERT genes with at least one of the studied disorders, twelve of them associated with two conditions. Gene-set analysis revealed significant associations with the DA gene sets for ADHD, ASD, MD and the cross-disorder GWAS meta-analysis, and with SERT gene sets for MD and BIP. The results obtained support a cross-disorder contribution of these two neurotransmitters systems in several psychiatric conditions.

## Data Availability

The results that we present are based on the analysis of publicly available genetic data (GWAS summary statistics) that can be found at the Psychiatric Genomics Consortium website.

https://www.med.unc.edu/pgc/

## ACKNOWLEDGMENTS

The authors thank the PGC ADHD, ANO, ASD, BIP, MD, OCD, SCZ, TS and cross-disorder working groups for making accessible the data from their studies.

## AUTHOR CONTRIBUTIONS

AR, NF-C and B.C conceived and designed the study. JC-D and BT obtained the data, performed the bioinformatic analyses and wrote the manuscript. JC-D, BT and NF-C prepared Figures and Tables. BC and NF-C coordinated the study and supervised the manuscript preparation. All authors contributed to and approved the final manuscript.

